# A pivotal study on patients’ selection for BRCA1 and BRCA2 mutations by different models in Libyan breast cancer women

**DOI:** 10.1101/2023.01.20.23284677

**Authors:** Eanas Elmaihub, Inas Alhudiri, Adam Elzagheid, Fakria Elfagi, Elham Hassen

## Abstract

**Introduction:** The BRCA mutation spectrum of familial breast cancer in Libya remains unknown. Several genetic models developed to predict the probability of BRCA1/2 mutations have not been applied in Libya, where the NCCN criteria are used for highly penetrating breast cancer susceptibility genes. This study aimed to predict BRCA1/2 mutation probability in familial breast cancer and eligibility for genetic testing by using BOADICEA and BRCAPRO models and NCCN criteria.

**Methods:** BRCA1/2 mutations were retrospectively predicted in 62 unrelated women with familial breast cancer between 2018 and 2021. Logistic regression, ROC analysis, and AUC were used to compare NCCN referral criteria with the BRCAPRO and BOADICEA scores.

**Results:** Of 62 breast cancer patients, 32 (51.6%) (mean age 43.5±8 years) were predicted by both models as BRCA mutation carriers. BRCAPRO predicted BRCA1 and BRCA2 mutations in 27.4% and 41.9% of the women, respectively. BOADICEA predicted 8% for BRCA1 and 29% for BRCA2. At least one NCCN criterion was met by 50/62 women (80.6%). Three criteria were statistically significant predictors in BRCAPRO and BOADICEA: breast cancer at ≤ 50 years with one or more close blood relatives with breast cancer, breast cancer patient with a close relative of male breast cancer, and triple-negative breast cancer. For the three respective criteria, sensitivity was 0.78, 0.89 and 0.75, specificity was 0.33, 0.39 and 0.22, AUC was 0.72, 0.75 and 0.76, PPV was 78%, 27.5% and 33.3, and NPV was 67%, 97% and 95.5.

**Conclusions:** BODICEA and BRCAPRO models are suitable for recommending genetic testing for BRCA gene mutations. The NCCN criteria are too broad.

## Introduction

Breast cancer (BC) has become the most common cancer worldwide. Although the incidence rates in transitioned countries are 88% higher than in transitional countries, the incidence rates in transitioned countries have increased rapidly [1]. North African countries are experiencing some of the most rapid increases, including Mauritania, Morocco, Algeria, Tunisia and Libya [1, 2]. In Libya, the incidence is 18.8 new cases per 100,000 women, most of the patients have advanced disease, and are often younger than in Europe, as is common in North Africa [3]. Although the rapid rise in BC incidence rates in North African countries could be attributed to lifestyle and environmental factors, the young age at onset and the high grade suggest the influence of genetic factors, such as mutations in breast cancer susceptibility genes (BRCA1 and BRCA2) [4]. The contribution of BRCA1/2 mutations to hereditary breast cancer has not yet been thoroughly investigated in North African populations [5, 6]. Hereditary genetic factors are responsible for 5–10% of all breast cancers, and BRCA1/2 gene mutations account for up to 25% of familial cases [7]. The lifetime cancer risk in BRCA1/2 mutation carriers is roughly 80–85% for female breast cancer (with older breast cancer onset for BRCA2), 12–40% for bilateral breast cancer for BRCA1, and 6–10% for male breast cancer [8–13]. These statistics are based on research on European and American populations, and they likely differ in other populations. This risk may be reduced by surgical interventions such as bilateral prophylactic mastectomy, which reduces the risk of BC by more than 90% [14–17]. Cancer susceptibility gene testing can identify individuals who are most likely to benefit from risk-reduction interventions.

Genetic testing of BRCA1 and BRCA2 is costly and difficult to interpret, particularly in families with a low likelihood of having a mutation. Therefore, it is important to estimate the likelihood that a family carries a BRCA1/2 gene mutation before a genetic test is done [18].

In several countries, a clinical standard for identifying hereditary breast/ovarian cancer has been developed and data have accumulated [15, 19]. This has resulted in the development of various models [20–23] that can more precisely estimate BRCA1/2 mutation carrier probabilities based on both genetic and empirical models that calculate the probability of mutation by using predictor variables derived from cancer family history. The BRCAPRO software [22, 24] and the Breast and Ovarian Analysis of Disease Incidence and Carrier Estimation Algorithm (BOADICEA) are examples of such tools [25, 26]. BOADICEA was founded by women and families from the United Kingdom, as well as a few other countries, and Africans are not represented. Furthermore, BRCAPRO is based on mutation rates and penetrance in women of Ashkenazi Jewish and European ancestry [8, 27].

Libya lacks genetic testing for familial and non-familial mutations in cancer patients, including breast cancer [28], and most of those who require it must travel abroad. Consequently, familial breast and ovarian cancer patients in Libya are assessed based on the National Comprehensive Cancer Network (NCCN) criteria for highly penetrating breast cancer susceptibility genes. In addition, the BRCA1/2 gene testing is typically recommended only for patients who are triple-negative and have a family history of breast/ovarian cancer. Despite the utility of the NCCN criteria for referral to genetic assessment, they lack specificity for accurately excluding people with low a priori risk [29]. In light of these issues, the logical step would be to develop local protocols that are efficient, practical, and beneficial to those specific patients.

To the best of our knowledge, BRCA1/2 mutation prediction models have not been applied to people of North African ethnicity. Hence, we aimed to retrospectively predict BRCA1/2 mutation probability in Libyan women with BC by using the BOADICEA and BRCAPRO models and to compare how the NCCN high-risk assessment criteria perform in referring BC patients for genetic evaluation in comparison to the two risk assessment models.

## Methods

### Patients and criteria

The study included 62 unrelated women with BC who were followed up at the outpatient clinic of the National Cancer Institute in Sabratha, Libya from 2018 to 2021. The patients were selected based on a family history of breast, ovarian, or other cancer in one or more first-second or third-degree relatives regardless of the age of onset. Data were collected from the patients in a self-administered questionnaire during their visits to the clinic. The medical records of the patients were reviewed.

### Study tools

To calculate the probability of a patient being a BRCA1/2 mutation carrier, we used the BRCAPRO CancerGene software program (v6, Bayes Mendel R package) (http://www4.utsouthwestern.edu/breasthealth/cagene) and BOADICEA CanRisk v6 (https://www.canrisk.org/).

BRCAPRO uses statistical ideas that go back to Bayes and Mendel [24]. It analyzes data from patients and all their relatives, including their age at diagnosis, current age, age at death, ethnicity, BC markers, such as the ER (estrogen receptor) scores, PR (progesterone receptor) and HER2 (human epidermal growth factor receptor-2), and other risk factors such as the woman’s age at first live birth, and mammographic density. The calculations make use of information on the occurrence of breast, ovarian, and other cancers among first– and second-degree relatives.

BOADICEA uses information such as lifestyle, women’s health, number and sex of children, breast screening, mammographic density, hormone receptors, including ER, PR, HER2, reproductive factors, and medical and family history. The medical history of the patients included age at diagnosis of breast or ovarian cancer. The family history included age at diagnosis of breast, ovarian, pancreatic, or prostate cancer in first or second-degree relatives. In our study, the ethnicity of all the patients was marked as “unknown” in BRCAPRO and as “other” in BOADICEA because of insufficient research on the ethnic groups living in North Africa. Based on the probability calculation scores in the two models, ≥ 10% was considered a high-risk subgroup that would benefit from genetic testing [30].

As reported in St. Gallen International Breast Cancer Conference 2011 [31], patients’ tumors were classified into four molecular subtypes according to the following definitions: luminal A (ER + and/or PR+, Ki67 low and HER2-), luminal B (ER + and/or PR+, Ki67 high and/or HER2+), HER2-positive (ER-, PR– and HER2+) and triple-negative (ER-, PR-, HER2-). Medical records were used to obtain ER, PR, HER2 and Ki67 expression levels. ER and PE were recorded in categories (positive vs. negative). Patients with l1 20% positive nuclei were identified as having high Ki67 expression, while those with ≤ 20% positive nuclei were identified as having low Ki67 expression. HER2 was considered positive when the score was +3 [32].

The patients were also assessed using the National Comprehensive Cancer Network NCCN criteria (v2.2022) for referral to genetic risk evaluation [33]. NCCN does not provide a percent probability of mutation, whereas the models calculate carrier probability scores for the BRCA1 and BRCA2 genes separately.

We compared each of the NCCN referral criteria to the BRCAPRO and BOADICEA scores of our breast cancer patients to determine how each of the NCCN criteria performed.

### Statistical analysis

We summarized the clinical characteristics of the study participants as medians for continuous variables and frequencies for categorical variables. Logistic regression was used in conjunction with receiver operating characteristic (ROC) analysis to determine which NCCN criteria were statistically significant predictors of high-risk patients, and the BRCAPRO and/or BOADICEA were used as the reference standards to evaluate the sensitivity and specificity of the patient’s criteria. The area under the curve (AUC), a general indicator of accuracy, was measured using ROC curves, and the calculations were made for both positive predictive values (PPV) and negative predictive values (NPV). A perfect test has an area of 1, whereas an area of 0.5 means that the test is useless. We examined each of the NCCN referral criteria concerning the BRCAPRO and BOADICEA results of our breast cancer patients to see how each of the NCCN criteria performed. The clinical significance of each NCCN criterion was determined by BRCAPRO and/or BOADICEA scoring. All statistical analyses were performed with IBM SPSS Statistics v20.

## Results

The clinical characteristics of the 62 BC women who participated in the study are summarized in Table 1. Forty-eight (77%) of them were ≤ 50 years old at diagnosis, with an average of 44.8 years. Thirty-eight women (61.2%) had a family history of BC in first-degree relatives and 13 (21%) in second-degree relatives.

**Table 1.**
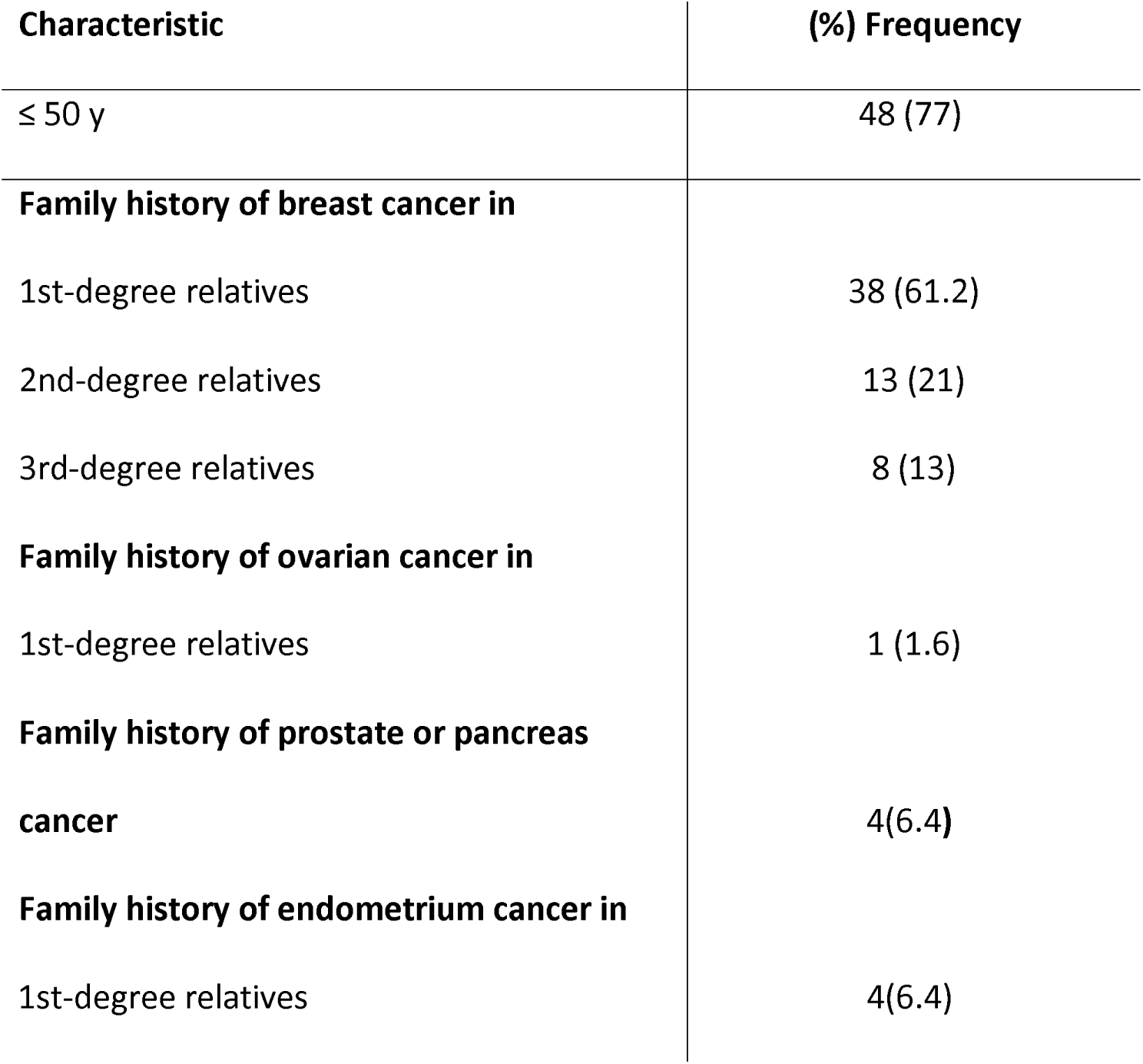
Patient characteristics.

In total, 62 patients were classified into four molecular subtypes of tumors (Table 2). More than half the patients were luminal B [38/62(61.3%)], followed by luminal A [11/62(17.7%)], triple-negative BC [8/62(13%)], and HER2+ [4/62(6.5%)]. Half of the BC patients aged ≤ 50 years had luminal B subtype (51.6%), whereas 11.3% had luminal A, 9.7% were triple-negative, and 4.8% were HER2 positive. Premenopausal and postmenopausal patients aged ≤ 50 years with the luminal A subtype represented 4.8% and 6.5%, respectively, while luminal B patients represented 38.7% and 12.9%.

**Table 2.**
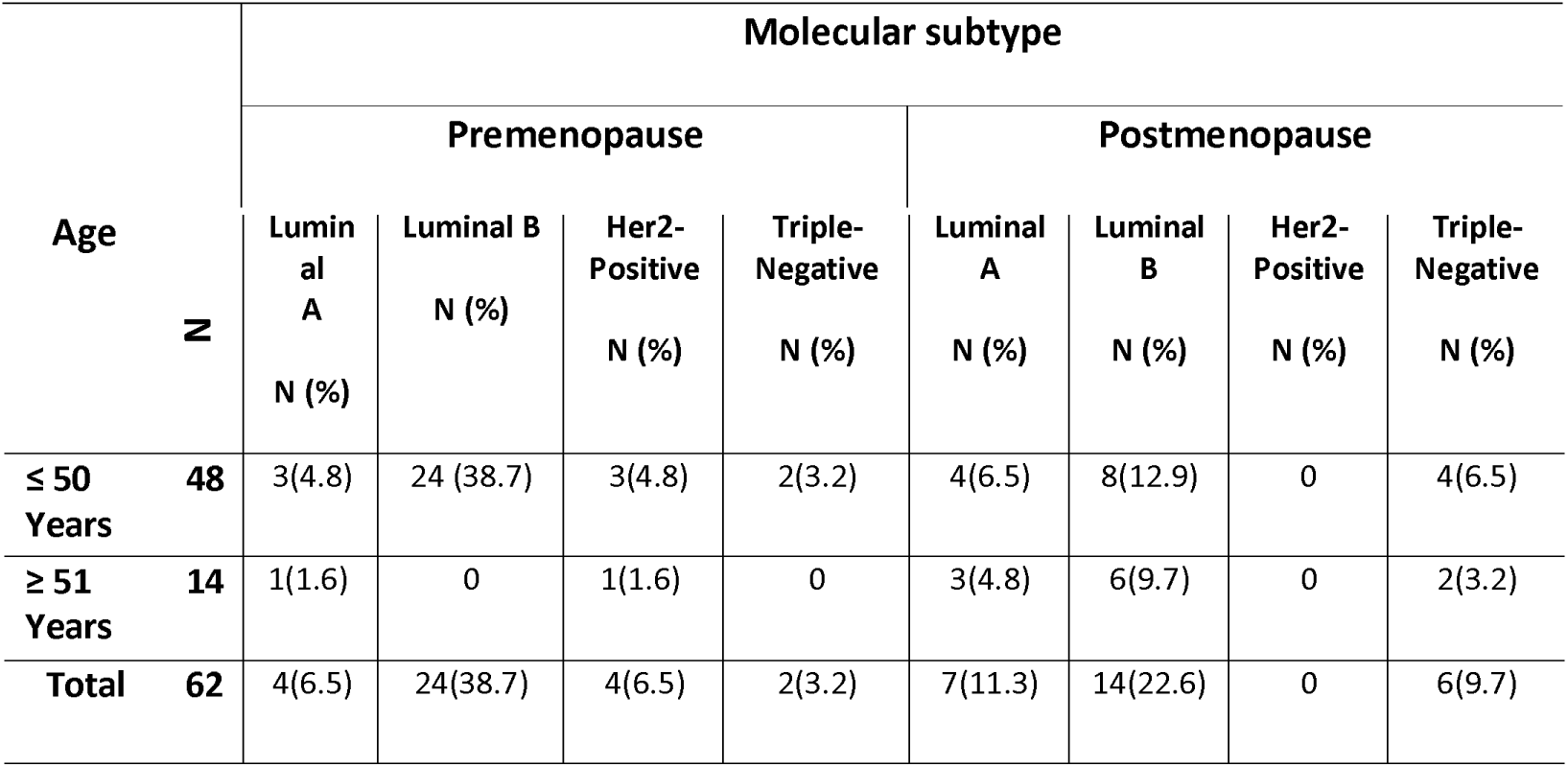
Distribution of molecular-subtype of breast cancer by age groups and menopausal status.

From the BRCAPRO and BOADICEA model scores based on the information provided by the patients, the highest prediction rates were 90.8% for BRCA2 and 75.7% for BRCA1. The patient with the highest BRCA2 score had a first-degree relative and a second-degree relative with a history of male breast cancer and was classified as a luminal B subtype. On the other hand, the highest score for BRCA1 was for a patient with double breast and ovarian cancer. The patient was classified as luminal A.

The mean carrier probability for all mutations in BRCAPRO was 8.1% for BRCA1 and 18.7% for BRCA2. For BOADICEA it was 4.8% for BRCA1 and 12.3% for BRCA2. Using a cutoff of ≥ 10%, BOADICEA identified 5/62 (8%) of the patients as having a high risk of BRCA1 mutations and 18/62 (29%) as high risk for BRCA2 mutations, whereas BRCAPRO identified 17/62 (27.4%) for BRCA1 and 26/62 (41.9%) for BRCA2. Consequently, 32/62 (51.6%) of the patients were identified as high risk by BOADICEA and/or BRCAPRO and therefore eligible for genetic testing. Four individuals had a high risk in both models of BRCA1, 13 had a high risk in BRCAPRO only, and 1 had a high risk in BOADICEA only. For BRCA2, 15 patients were positive in both models at the same threshold, 11 were positive in BRCAPRO, and 3 were positive in BOADICEA.

The average ages (mean ± SD) of patients classified as high risk for BRCA1 mutations by BRCAPRO and BOADICEA were, respectively, 42.5 ± 9.5 years and 43.6 + 14.8 years. For both models together, it was 42.7 ± 9.2 years. The mean age of patients identified as having a low risk of BRCA1 mutations by these models was 45.1 ± 8.9 years (p = 0.7). On the other hand, the average ages of patients with a high risk of BRCA2 mutation by BRCAPRO, BOADICEA, both models and low-risk BRCA2 mutations were 42.12 ± 7.8 years, 43.5 ± 9.6, 43.6 ± 8.4 and 45.8 ± 9.2, respectively (p = 0.8).

Table 3 shows the results for 50/62 (80.6%) patients who met at least one NCCN criterion. Using logistic regression, NCCN criterion (BC patient at age ≤ 50 with one or more BC close blood relatives) was a statistically significant predictor of patients identified as high risk for BRCA1 and BRCA2 2 mutations by either BRCAPRO or BOADICEA score (p = .011 and p = .005 respectively) in with sensitivity of 0.78, specificity of 0.33, and area under the ROC curve of 0.72 (Fig 1). PPV was 78% and NPV was 67%. The criterion of BC patient with relative male BC was statistically significant in patients at high risk of BRCA2 mutations (P = 0.023), with a sensitivity of 0.89, specificity of 0.39, and area under the ROC curve of 0.75 (Fig 2). PPV was 27.5 % and NPV was 97%. Triple-negative BC patients diagnosed at age ≤ 60 were also statistical significant at high risk patients of BRCA1 mutations (P = 0.008), with a sensitivity of 0.75, specificity of 0.22 and area under the ROC curve of 0.76 (Fig 3). PPV was 33.3% and NPV 95.5%.

**Fig 1.**
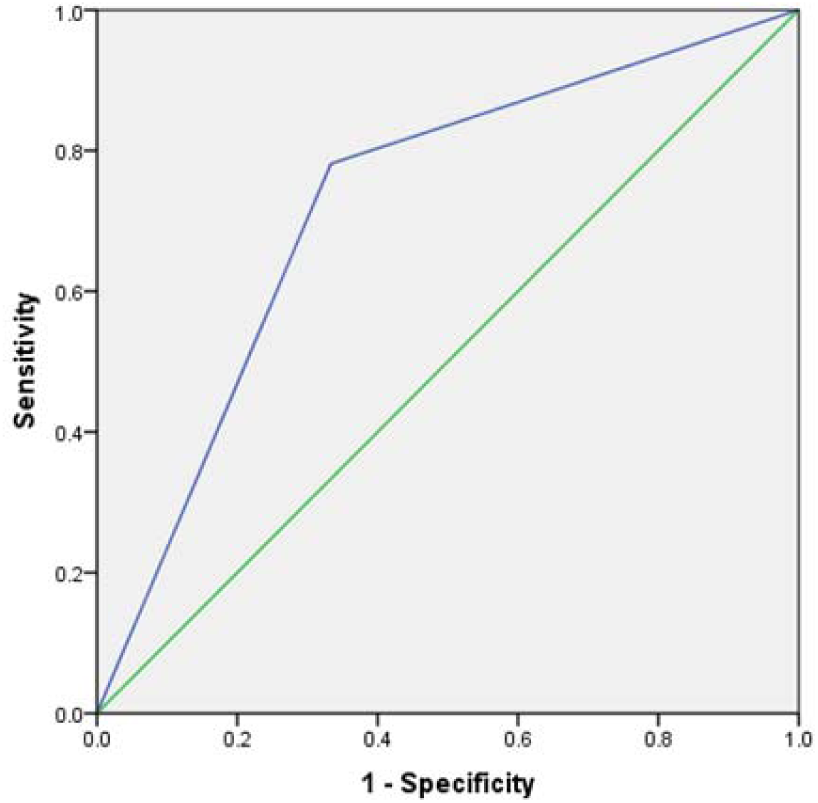
ROC curve for NCCN criterion of BC at age ≤ 50 years and one or more close blood relatives with breast or ovarian cancer at age ≤ 50 years as a predictor for genetic test eligibility.

**Fig 2.**
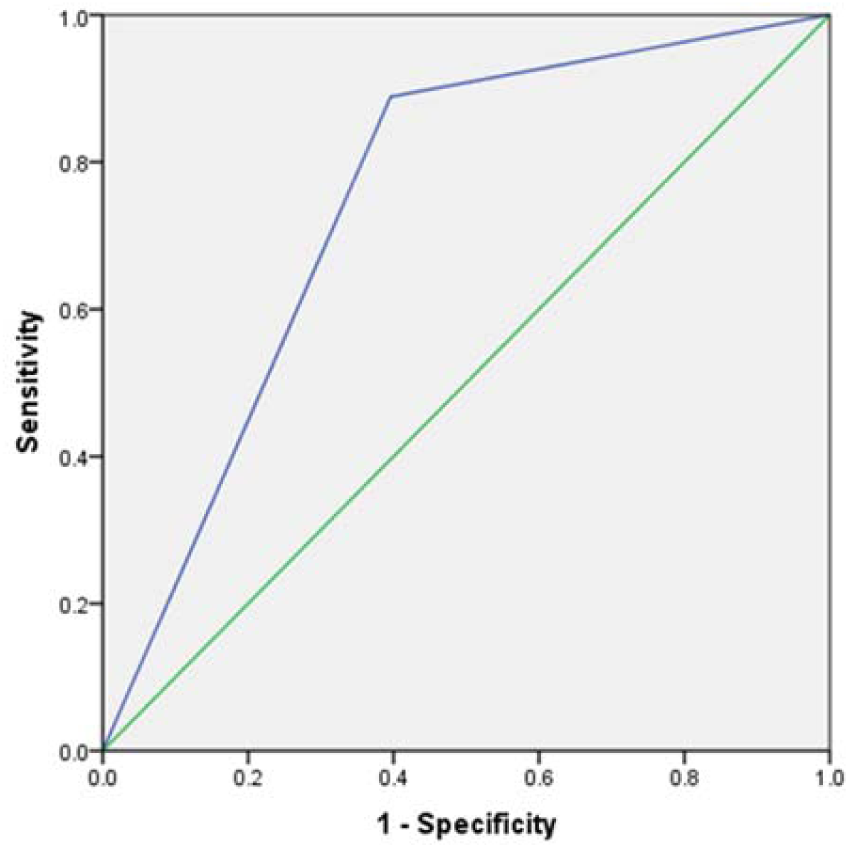
ROC curve for NCCN criterion: BC patients with relatives of male BC as a predictor of genetic test eligibility.

**Fig 3.**
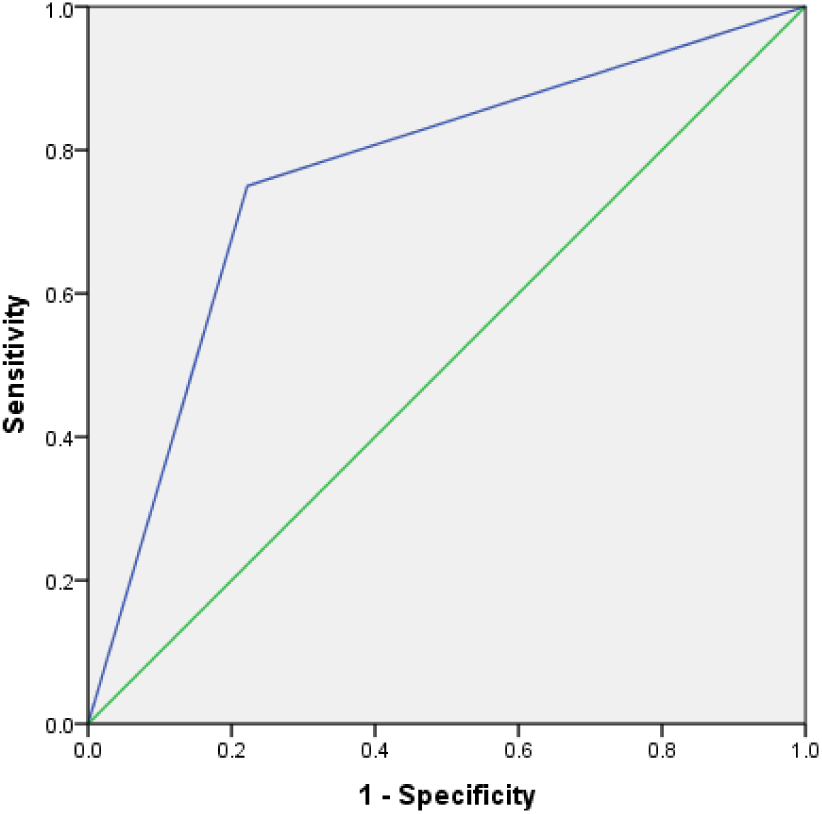
ROC curve for NCCN criterion: Triple-negative (ER, PR, Her2-) BC diagnosed ≤60 years old as a predictor of genetic test eligibility.

**Table 3.** BRCAPRO and BODICEA models against NCCN criteria for BRCA1/2 genes mutation risk prediction.

| NCCN Criteria<br>A woman with<br>BC diagnosis<br>meeting any of<br>the following | No. of patients | BRCAPRO<br>patients score ≥<br>10 | | BODICEA<br>patients<br>score ≥ 10 | | High score by<br>BRCAPRO<br>or BOADICEA | | P value for the<br>high score by<br>BRCAPRO or<br>BOADICEA | |
| --- | --- | --- | --- | --- | --- | --- | --- | --- | --- |
|  |  | BRCA1<br>N = 17 | BRCA2<br>N = 26 | BRCA1<br>N = 5 | BRCA2<br>N = 18 | BRCA1<br>N = 18 | BRCA2<br>N = 29 | BRCA1 | BRCA2 |
| BC at age ≤ 50<br>and ≥ 1 close<br>blood relative<br>with B or O C<br>at age ≤ 50 | 35 | 14 | 22 | 4 | 14 | 15 | 23 | .011 * | .005 * |
| At age $45 \leq$ | 10 | 1 | 1 | 0 | 0 | 1 | 1 | .178 | .031 |
| Double<br>primary BC/<br>OC | 2 | 1 | 2 | 1 | 1 | 1 | 2 | .487 | .999 |
| Male BC<br>relative | 9 | 4 | 9 | 0 | 6 | 4 | 9 | .279 | .023 * |
| ≥ 1 relative<br>with prostate<br>or pancreatic<br>cancer | 4 | 0 | 1 | 0 | 1 | 0 | 1 | .999 | .385 |
| Triple-negative<br>(ER, PR, Her2-)<br>BC diagnosed<br>≤60 years old | 8 | 5 | 3 | 1 | 3 | 6 | 4 | .008 * | .845 |

Twelve patients did not meet any of the NCCN criteria.
Breast B, Ovary O, Cancer C, \* Significant.

## Discussion

Despite recent efforts by Libyan health authorities to develop nationwide breast cancer screening programs [34], the unavailability of genetic diagnostics for people with familial breast and ovarian cancer remains a concern. Situation of BRCA mutations in Libyan women has not been practiced, and this is the first study to use the BRCAPRO and BOADICEA scoring (≥ 10%) to predict BRCA1/2 gene mutations in women with BC who have a strong family history of BC and to evaluate the efficacy of NCCN criteria.

Based on the BRCAPRO and BOADICEA scoring systems, our calculated frequency of patients at high risk of being carriers of BRCA1 and BRCA2 mutations were 29% and 46.8% for the respective scoring systems. If these high-risk patients were to test positive for BRCA1/2 mutations, this estimate would be consistent with studies showing that BRCA2 mutations are more common than BRCA1 mutations in the Arab region [35], though this pattern is not observed in the majority of other populations [36]. We also found that the occurrence of both breast and ovarian cancer in one patient was related to a high risk of a BRCA1 mutation, as reported in Sweden [37]. This had the highest probability of BRCA1 mutation in the BODICEA model.

We employed BRCAPRO and BOADICEA because they are the most reliable in predicting mutant carrier probability when compared to other scoring models [38]. BRCAPRO predicted that 27.4% of the patients had BRCA1 mutations and 41.9% had BRCA2 mutations, which are slightly greater than the percentages predicted by BOADICEA (8% for BRCA1 and 29% for BRCA2). The BRCAPRO model is similar to BOADICEA in that BRCA1 and BRCA2 are modeled independently. The differences between them can be explained in part by their use of different mutation rates and allele frequencies. In particular, in BRCAPRO, the mutations are assumed to be more common in BRCA1 than in BRCA2, but BOADICEA finds that BRCA1 and BRCA2 mutations have similar population frequencies, though BRCA2 was more common [22]. The benefit of using two predictive models instead of one was that we were able to identify more high-risk patients who needed genetic testing, and based on score results from either or both scoring systems, we recorded 13 more BRCA1 and 11 more BRCA2 high-risk patients compared to using either system alone.

Patients with BC at the age of ≤ 50 and having first or second-degree relatives with BC at the age of ≤ 50 met the highest risk NCCN criteria. The average age of these patients was 44.8 years, which is consistent with a previous study in Libya that showed a higher frequency of BC among those ≤ 50 years old [3, 39-41]. Luminal B subtype (61.3%) was more prevalent than luminal A (17.7%). This is in agreement with studies in Tunisia [42], Morocco [43], Saudi Arabia [44] and Italy [45] but differs from studies in other North African countries, including Egypt, Tunisia and Algeria [46–49]. The difference could be explained by the heterogeneity of BC in different cohorts drawn from different countries. The incorporation of HER2-positive patients in luminal B groups and the use of different Ki-67 cut-off values to stratify luminal breast cancer groups around the globe could lead to different results [42]. Furthermore, the 13% triple-negative breast tumors in our study is similar to the percentages found in Tunisia and Morocco [42–43], and the 6.5% for the Her2-positive subtype is in agreement with studies from Algeria, Europe and the USA [50].

Several studies refer to a link between specific molecular subtypes and BRCA1/2 mutation status. BRCA1/2 mutation carriers are more likely to have triple-negative BC. This association is strongest in BRCA1-related BC, and most BRCA2 BCs belong to the luminal B subtype [51]. Interestingly, the results of our study show that 75% of the triple-negative BC patients had scores ≥ 10 of carrier BRCA1 mutation, and 55.3% of the luminal B subtype had BRCA2 scores ≥ 10. These findings need confirmation by future full gene sequence analyses for BRCA1 and BRCA2 in the same patients.

The overall mutation prevalence in patients identified as high risk by BRCAPRO and BOADICEA was lower than would be expected based on the NCCN criteria. The NCCN criteria that correlated best with the scoring systems for identifying high-risk patients were as follows: (i) BC patients at age ≤ 50 years and having one or more close blood relatives with BC at age ≤ 50 years, (ii) BC patients having male relatives with BC, and (iii) patients with triple-negative BC (ER, PR, Her2) diagnosed at age ≤ 60 years. For these, sensitivity was high (78%, 89% and 75%, respectively) and specificity was low (33%, 39% and 22%, respectively). This could result in a large number of patients with a low likelihood of having a BRCA mutation being referred for a genetic test. Though the P value of BC patients with BRCA2 at age ≤ 45 years was .032 it had a poor area under the ROC curve (0.4), indicating that the criterion may not differentiate between high– and low-risk BRCA mutation carriers, especially because it depends on patient age alone. The other criteria (double primary BC and one or more relatives with prostate or pancreatic cancer,) were not significant, possibly due to the small sample size.

Our study has some limitations. We hypothesized that a BRCAPRO or BOADICEA cutoff of ≥ 10% would be enough to predict the presence of a BRCA mutation in patients who require a genetic test with acceptable false-negative rates. We also could not compensate for the fact that none of our patients had a genetic test. However, this study aims to make a pretest prediction of BRCA1 and BRCA2 carrier mutations. We also had only one patient with double primary BC, one with double primary BOC, four BC patients with relatives who had prostate or pancreatic cancer, and eight triple-negative (ER, PR, Her2) BC patients at the age ≤ 60. As a result, increasing the number of patients who meet the rarer criteria will improve the statistical analysis of that criterion.

## Conclusion

The NCCN criteria for making decisions on genetic testing have shortcomings because some of them are too general. Based on our results, we believe that using BRCA risk calculator models such as BRCAPRO and BODICEA will be advantageous for identifying high-risk patients. This pivotal study’s results will be validated by performing BRCA1/2 testing based on the BRCAPRO and BOADICEA models.

## Declarations

### Ethics approval and consent to participate

The study was approved by the Research Ethics Committee of the National Cancer Institute (NCI), Sabratha, Libya. Verbal informed consent was obtained from each patient before recruitment in the study, which was approved by the Research Ethics Committee of the National Cancer Institute (NCI), Sabratha, Libya. This study was conducted in accordance with the ethical principles of the Helsinki Declaration.

### Consent for publication

Not applicable.

### Availability of data and materials

All data generated or analyzed during this study are included in supplementary information files.

### Author contributions

Conceptualization: Eanas Elmaihub, Inas Alhudiri

Data curation: Eanas Elmaihub, Fakria Elfagi

Formal analysis: Eanas Elmaihub

Investigation: Eanas Elmaihub, Fakria Elfagi

Methodology: Eanas Elmaihub, Inas Alhudiri

Supervision: Elham Hassen, Adam Elzagheid

Writing – original draft: Eanas Elmaihub

Writing – review & editing: Elham Hassen, Inas Alhudiri

## Data Availability

All data produced in the present study are available upon reasonable request to the authors

## Acknowledgment

We appreciate the help received from the Sabratha Research Unit of the National Cancer Institute and the Oncology Department.

